# Maternal Extracellular Vesicles During Pregnancy and Autism Risk in Children

**DOI:** 10.64898/2026.06.25.26355526

**Authors:** Delia McGowan, Serena Nencini, William Yakah, Claire-Marie Vacher, Helene Lacaille, David M. Haas, William A. Grobman, Robert M. Silver, Uma M. Reddy, Ronald J. Wapner, William P. Fifer, Anna A. Penn, Morgan R. Firestein

## Abstract

**Background:** Differences in extracellular vesicles (EVs), bioactive nanoparticles involved in intercellular signaling, have been reported in those with autism. However, little is known about the association between maternal EVs during pregnancy and the likelihood of autism in offspring. This study evaluated the association of the concentration and cargo material of EVs in prenatal maternal plasma with childhood autism likelihood.

**Methods:** Participants in the Nulliparous Pregnancy Outcomes Study provided maternal plasma at 15–23 weeks’ gestational age. EVs were isolated by ultracentrifugation, and concentration, mean size, CD63 levels, and RNA cargo were assessed by nanoparticle tracking analysis, ELISA, and small RNA sequencing. At 4.5–6 years of age, parents completed the Social Communication Questionnaire. Thirty-one children at high-risk for autism were matched to 31 low-risk children on sex, age, and gestational age. Differential RNA transcript analysis and over representation analysis were performed.

**Results:** There were no group differences in CD63 levels, mean particle size, or EV concentration (*p*>0.1). Nominal bin-level differences were observed at 280–290 nm and 430–440 nm before multiple-comparison correction. One hundred forty-five RNAs, including protein-coding RNAs, piRNAs, lncRNAs, miRNAs, snoRNAs, snRNAs, and tRNAs, were differentially contained, most of them downregulated in those at high risk of autism. These RNAs mapped to pathways involved in immune/inflammatory signaling, intracellular trafficking, protein turnover, and neurodevelopment. Six of the 62 (9.7%) differentially contained protein-coding RNAs overlapped with genes in the SFARI Gene database.

**Limitations:** Large studies involving individuals diagnosed with autism are needed to evaluate the role of prenatal EVs in the pathogenesis of the condition. Additionally, prenatal sampling of EVs across multiple timepoints and subsequent deconvolution to determine the source of the EVs will strengthen interpretability and veracity of our findings.

**Conclusions:** These findings provide preliminary evidence that maternal prenatal EV RNA cargo is associated with childhood autism likelihood.

## Introduction

Despite the prevalence of autism, the underlying biological mechanisms remain elusive and diagnostic biomarkers are limited. Autism is a heterogeneous, neurodevelopmental condition typically diagnosed during early childhood and characterized by social communication difficulties and restricted, repetitive patterns of behaviors and interests^1^. Current screening and diagnostic tools rely on observable behaviors that do not emerge until toddlerhood or later, delaying access to services^2,3^. Therefore, early-emerging biological indicators of autism likelihood hold the potential to both expedite access to early intervention services and further our understanding of autism etiology. Extracellular vesicles (EVs) are emerging as a potential biological mechanism, marker, and therapeutic platform for psychiatric conditions, including autism^4–6^.

Exosomes and larger EVs are lipid bilayer-enclosed particles that are naturally released from mammalian cells and are carried through the bloodstream to recipient cells^7,8^. EVs carry proteins, lipids, and RNAs that mediate intercellular communication and cellular processes^8–10^. They can be readily isolated from most human biofluids^11–13^, making their utility as a non-invasive biomarker particularly promising^14–17^. Distinct and overlapping EV-contained microRNAs (miRNAs) have been identified in plasma and brain-derived EVs from individuals with depression, bipolar disorder, and schizophrenia, suggesting that EV cargo may reveal convergent processes that are disrupted across psychiatric conditions while providing insight into specialized biomarkers for individual conditions^18,19^.

In autism, studies have reported differences in EV-associated protein levels, particle concentration, RNA cargo, and protein cargo assessed in pediatric and adult populations^20–25^. Although findings regarding EV size and concentration have been mixed, studies of plasma, neuron-derived, and organoid-derived EVs consistently suggest that EV cargo is altered in autism, converging on neuronal, cytoskeletal, focal adhesion, signal transduction, axon guidance, immune, coagulation-related, glycan-related, and metabolic pathways^21–24^. These differences may emerge early in development, as umbilical cord blood EVs from infants later diagnosed with autism exhibit distinct proteomic profiles^25^. While less is known regarding the downstream actions of EVs following cellular uptake, EVs from the serum of autistic children induce cultured human microglia to secrete higher levels of IL-1β^20^, suggesting the EVs may contribute to the dysregulated microglial activity and elevated inflammatory cytokine levels reported in subpopulations of autistic individuals^25^.

Autism likely manifests during fetal development, and mounting evidence implicates several pregnancy-related factors in the pathogenesis of later psychopathology^27,28^; however, the association between EVs during pregnancy – including those of maternal, fetal, and placental origin – and autism in offspring remains largely unexplored. Beyond their role in cell-to-cell communication, EVs are involved in maternal-fetal-placental communication^29–31^. Throughout pregnancy, the maternal and fetal systems are connected through placental syncytiotrophoblast cells (STBs)^9,32^, which form the primary interface between mother and fetus and are the primary source of placenta-derived EVs^32,33^. Fluorescently labeled EVs of fetal origin have been detected in maternal blood, and bidirectional flow of EVs across the placenta have been observed using bioengineered EVs^34^. Placental EVs can be detected in maternal blood by six weeks of gestation and account for ∼10-20% of the EVs in maternal blood during pregnancy^31^. The concentration and cargo of EVs are altered in pregnancies complicated by conditions associated with increased autism risk, including gestational diabetes mellitus (GDM) and hypertensive disorders of pregnancy. In women with GDM, placental exosomes increase twofold compared to women without GDM^35,36^, and exhibit modified bioactivity, including increased release of pro-inflammatory cytokines^36^, increased levels of dipeptidyl peptidase IV^37^, and differential expression of seven miRNAs^38,39^.

Taken together, EVs are a compelling candidate mechanism through which prenatal exposures may contribute to neurodevelopment and autism likelihood. Whether EVs in maternal plasma during pregnancy relate to autism in children remains largely unknown. The present study represents convergence of ongoing, but separate lines of research to examine the association of maternal EVs during pregnancy with subsequent autism risk in children.

## Methods & Materials

### Study Participants

Participants previously enrolled in the multi-site Nulliparous Pregnancy Outcomes Study: Monitoring Mothers-to-be (nuMoM2b). Between 2010-2013, the nuMoM2b study enrolled 10,037 pregnant individuals in the first trimester of pregnancy across 8 clinical sites, including Columbia University Irving Medical Center (CUIMC). Inclusion and exclusion criteria have been described previously^40^. This analysis includes data and biospecimens from a subset of the 1,005 participants who were enrolled at the CUIMC site. Through a previous ancillary study^41^ of 661 participants who provided consent for future use of data, future contact by study staff, and maternal plasma during the second trimester, mothers were invited to completed the parent-reported Social Communication Questionnaire^42^ (SCQ) to assess autism behaviors at age 4.5-6 years.

In total, 270 mothers completed the SCQ through an online data capture portal. Using a pre-defined total score cutoff of 10^43^, we identified 31 full-term children as being at high risk for autism. These 31 children were matched on sex, gestational age at birth, and age at the assessment with 31 children at low risk based on SCQ total scores, resulting in a sample size of 62 children. Since the sample size was limited to the number of respondents and the rate of positive autism screenings in the sample, analyses are considered exploratory.

### Social Communication Questionnaire (SCQ)

At age 4.5-6 years (mean=63.7 months, range=55.0-75.0 months) mothers completed the lifetime version of the SCQ^42^, a parent-report questionnaire to evaluate autism related behaviors in children 4 years and older. The SCQ contains 40 yes-or-no items to determine whether the child exhibits behaviors associated with autism. The SCQ provides a continuous score, with higher scores indicating more autistic behaviors. Children with scores equal to or above a pre-established cutoff of 10 were considered to be at high-risk^43^.

### Maternal Plasma Collection

Whole blood was collected by venipuncture into K2EDTA-coated tubes between 15 and 23 weeks of gestation (mean: 19.2±1.3 weeks). Collection tubes were inverted to distribute the K2EDTA and centrifuged at 1500g for 10 min at 4 °C. Plasma was stored at -80 °C until further processing, and all samples underwent one freeze-thaw cycle prior to EV isolation.

### EV Isolation

EVs were isolated from maternal plasma. Variable amounts of plasma were available across samples. Therefore, plasma was diluted with sterile filtered phosphate-buffered saline (PBS, pH 7.4) as needed to reach a volume of 1 mL. Samples were subjected to two rounds of ultracentrifugation at 25,000 × g and 4°C (2h followed by 1h). After the first spin, the supernatant was removed and the EV pellet was resuspended in PBS prior to the second ultracentrifugation. Final EV pellets were resuspended in sterile PBS (450 μL) and stored at −80°C.

### Quantification of CD63 by ELISA

Levels of the exosome membrane marker CD63 were quantified using the Human CD63 ELISA Kit (ab275099, Abcam, Cambridge, UK) according to the manufacturer’s instructions. Briefly, a volume of 50 µL of each sample or standard was added in duplicate to the pre-coated microplate wells and incubated for 1 hour at room temperature on a plate shaker set to 400 rpm, with 50 µL of Antibody Cocktail added to each well. The colorimetric reaction was developed by adding TMB substrate and incubating for 20 minutes at room temperature in the dark on a plate shaker. The reaction was stopped using the provided stop solution. Absorbance was measured at 450 nm using a microplate spectrophotometer. CD63 concentrations were interpolated from a standard curve and expressed as ng/mL. Results were adjusted according to the plasma dilution factor.

### Nanoparticle Tracking Analysis

Nanoparticle tracking analysis (NTA) was performed using the Horiba ViewSizer 3000 to quantify the size distribution and concentration of EVs. EVs were diluted to 1:500 with PBS. The concentration of EVs was determined for particles ranging from 30-500 nm and was adjusted for the plasma dilution factor.

### Transmission Electron Microscopy

Isolated EVs were assessed by transmission electron microscopy (TEM). Briefly, 5 μL of the diluted EVs suspended in PBS were deposited on a Formvar/Carbon 200 Mesh, Copper electron microscopy grids (Electron Microscopy Sciences, #FCF200-Cu-50) and left to adsorb for 1 min. The grids were then washed two times with 10 μL of distilled water for 1 min. The grids were then stained with Stain77 (Electron Microscopy Sciences, #22409) for 1 min. After drying for 5 min at room temperature, grids were examined in the Philips CM12 transmission electron microscope at 80 Kv.

### Small RNA Sequencing of EV Cargo

RNA was isolated from EVs using the Qiagen miRNeasy Serum/Plasma Advanced Kit and total RNA concentration was determined. Small RNA sequencing was performed by RealSeq Biosciences (Santa Cruz, CA, USA). Half-volume RealSeq-Biofluids Small RNA libraries were prepared using 6 μL of RNA. The libraries were pooled to equal nanomolarity concentrations and purified and size-selected using Pippin Prep (Sage Biosciences). The library pools were profiled using DNA Tapestation and dsDNA High Sensitivity Qubit before sequencing on the Singular Genomics G4. Sequencing was performed with single 100bp reads. Raw fastq files were processed using *Cutadapt*. Adapter sequences were removed and reads were filtered based on length. Reads shorter than 1 bp were filtered to determine the amount of adapter dimer and reads with a minimum 15 bp length were aligned to a reference. Trimmed reads were mapped to various databases sequentially with bowtie2. Raw counts of 25,725 RNAs were processed and filtered to remove mitochondrial RNAs (n=8,514) and RNAs with zero counts across all samples or for which ≥25% of the samples had a count of zero (n=8,490), resulting in 8,721 RNAs in the differential expression analysis. The list of differentially EV-contained protein-coding RNAs generated by DESeq2 analysis was further analyzed using the enrichR package in R^44–46^. Over representation analysis identified significantly enriched biological processes using the Kyoto Encyclopedia of Genes and Genomes (KEGG) 2026 and Gene Ontology (GO) Biological Process 2025 databases, and we further analyzed for enrichment of autism associated genes reported in the SFARI Gene database^47^, which catalogs genes associated with autism and provides evidence-based scores reflecting the strength of each gene’s association with autism.

## Statistical Analysis

Statistical analyses were performed in R version 4.5.2 (2021-11-01) and Prism 10. Scores on the SCQ were analyzed as both a binary and continuous variable. We fit generalized linear models (GLMs) with a Gaussian distribution to assess associations of EV concentrations with continuous SCQ total scores, and GLMs with binomial distribution and logit link (logistic regression) or Welch’s two-sample T-tests to evaluate associations of EV concentrations with binary SCQ risk status. Primary analyses were conducted in the overall sample and separate GLMs were fit for male and female children to estimate sex-specific associations. The SCQ groups (high risk versus low risk) were matched based on infant sex assigned at birth, infant gestational age at birth (± 5 weeks), and child age at the assessment (± 7 weeks). To determine whether the models required further adjustment, we performed Pearson correlation tests and Welch’s two-sided t-tests to identify which, if any, of the potential covariates (child’s sex, gestational age at birth, age at assessment, maternal age at delivery, maternal ethnicity) were statistically significantly associated with either the dependent or independent variables. None of the associations reached statistical significance (p>0.05). To compare group differences in EV concentration across particle sizes, we performed multiple Welch’s two-sample T-tests with Bonferroni correction. The DESeq2 R package was used to identify differentially contained RNA transcripts (*p*<0.01). Over representation analysis was performed, and KEGG and GO Biological Processes pathways with at least three contributing genes and a p-value of <0.1 were determined. For all other analyses, statistical significance was set to alpha=0.05.

## Results

### Participants

Demographic and clinical characteristics of participants for each SCQ group are provided in **Table 1**. The average maternal age across both groups was 34.2 years (range 26-47 years) and the average paternal age was 30.6 years (range 18-48 years). 55% and 65% of mothers self-identified as Hispanic or Latino in the low-risk and high-risk SCQ groups, respectively. In the overall sample and across both groups, most mothers completed a graduate or professional degree. The sample was matched based on child’s sex assigned at birth. Therefore, the groups include equal numbers of females (n=28) and males (n=34). The average gestational age at birth was 39 weeks±3 days (range 37 weeks±0 days – 40 weeks±0 days), the average birth weight was 3320 grams (range 2130-4550 grams), and 16 (26%) of the infants were delivered by cesarean delivery. The groups did not differ significantly regarding baseline demographic or clinical characteristics (**Table 1**).

**Table 1.** Demographic and Clinical Characteristics of the Study Sample.

| | Low-Risk<br>N=31<br>Mean (SD);<br>No. (%) | High-Risk<br>N=31<br>Mean (SD);<br>No. (%) | Test statistic<br>t-test or $\chi^2$ | p-value |
| --- | --- | --- | --- | --- |
| **Parental Characteristics** |  |  |  |  |
| Maternal Age (years) | 34.8 (5.3) | 33.6 (6.3) | 0.80 | 0.425 |
| Unknown | 3 | 3 |  |  |
| Paternal Age (years) | 30 (6) | 31 (8) | -0.71 | 0.481 |
| Unknown | 1 | 0 |  |  |
| Maternal Educational Attainment |  |  | 2.6 | 0.625 |
| No high school degree | 1 (3.2%) | 0 (0%) |  |  |
| High school degree | 4 (13%) | 7 (23%) |  |  |
| Some college, no degree | 4 (13%) | 5 (16%) |  |  |
| college degree | 7 (23%) | 8 (26%) |  |  |
| Graduate/professional degree | 15 (48%) | 11 (35%) |  |  |
| Maternal Ethnicity |  |  | 0.27 | 0.605 |
| Not Hispanic/Latino | 14 (45%) | 11 (35%) |  |  |
| Hispanic/Latino | 17 (55%) | 20 (65%) |  |  |
| Mode of Delivery |  |  | 0.08 | 0.772 |
| Cesarean | 9 (29%) | 7 (23%) |  |  |
| Vaginal | 22 (71%) | 24 (77%) |  |  |
| **Infant Characteristics** |  |  |  |  |
| Sex |  |  | 0.00 | >0.999 |
| Female | 14 (45%) | 14 (45%) |  |  |
| Male | 17 (55%) | 17 (55%) |  |  |
| Birth Weight (grams) | 3,375 (490) | 3,266 (453) | 0.91 | 0.365 |
| Head Circumference (cm) | 34.25 (1.37) | 33.94 (1.70) | 0.80 | 0.428 |
| Unknown | 1 | 0 |  |  |
| Gestational Age at Birth (weeks) |  |  | 1.8 | 0.780 |
| 37 | 2 (6.5%) | 2 (6.5%) |  |  |
| 38 | 5 (16%) | 8 (26%) |  |  |
| 39 | 13 (42%) | 13 (42%) |  |  |
| 40 | 8 (26%) | 7 (23%) |  |  |
| 41 | 3 (9.7%) | 1 (3.2%) |  |  |
| Age at Follow-Up (months) | 64.1 (6.1) | 63.2 (5.9) | 0.60 | 0.554 |

Children in the high-risk group had total SCQ scores that were ≥10, with a mean of 13.97 (range 10-32). Children in the low-risk group had a mean total score of 3.39 (range 0-9). The groups differed significantly on total SCQ scores (*t*(43)=-11.61, *p*<0.001).

### Characterization of EVs

Representative TEM images of EVs isolated from maternal plasma showed membrane-bound vesicles with round morphology and sizes consistent with EVs (**Figure 1A**). The mean CD63 concentrations of EVs in plasma of mothers of children with low versus high SCQ scores were 496pg/mL and 720pg/mL, respectively. The CD63 concentration did not differ significantly between the groups (*t*(47.98)=-0.80, *p*>0.1) (**Figure 1B**). Adjusted and unadjusted GLMs further demonstrated no significant association of CD63 with total SCQ scores. Similarly, there were no group differences either with regard to the concentration of EVs as determined by NTA (low-risk mean=3.422e+07, high-risk mean=3.292e+07; *t*(59.96)=-0.28, *p*>0.1) (**Figure 1C**) or in the mean size of EVs (low-risk mean=120.48, high-risk mean=122.32; *t*(58.81)=-0.58, *p*>0.1) (**Figure 1D**). To determine whether EV concentration differed between groups at specific particle sizes, the size distribution was aggregated into 10-nm bins. For each bin, EV concentration was summarized at the participant level and group differences were assessed using Welch’s two-sample t-tests and the Benjamini–Hochberg false discovery rate (FDR) procedure was applied. A nominal difference was observed at 280-290 nm and at 430-440 nm in the full sample and in women carrying male fetuses (**Figure 1E**); however, these differences did not survive correction for multiple comparisons.

**Figure 1.**
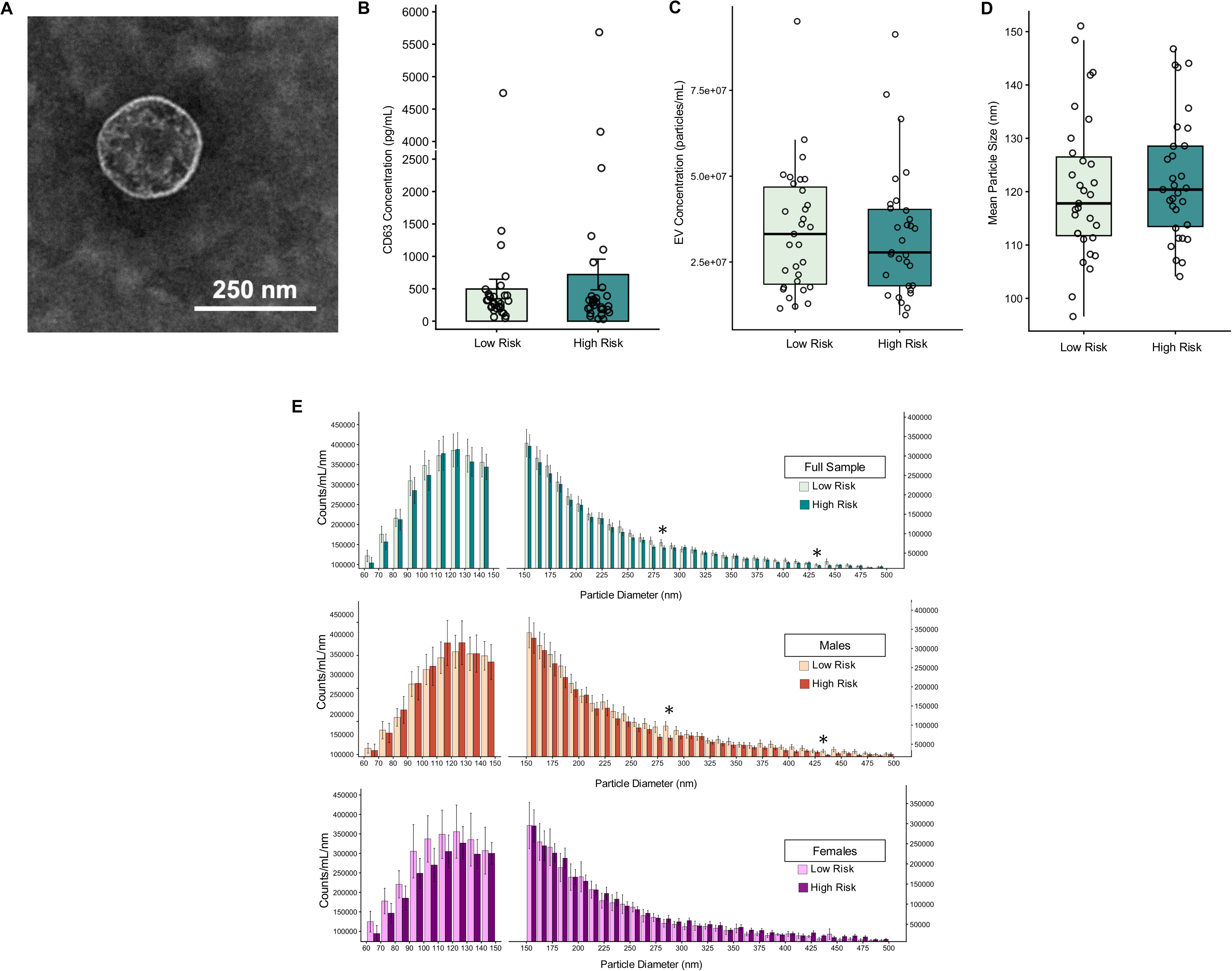
Characterization and concentration of EVs isolated from second trimester maternal plasma and childhood scores on the SCQ. (A) Transmission electron microscopy image of a small extracellular vesicle (exosome) isolated from maternal plasma. (B) CD63 concentration in EVs from maternal plasma from mothers of children at low risk versus high risk based on total SCQ scores at age 4.5-6 years. Error bars represent SEM. (C) Mean EV concentration as determined by nanoparticle tracking analysis in maternal plasma from mothers of children at low risk versus high risk based on total SCQ scores. (D) Mean particle size in maternal plasma from mothers of children at low risk versus high risk based on total SCQ scores. (E) A comparison of the distribution of EVs across 10 nm bins between children at low risk versus high risk in the full sample, among male children, and among female children at age 4.5-6 years. Mean ± SEM; An asterisk denotes a significant group difference of *p*<0.05.

### Small RNA-seq Analysis of EV Cargo

In the full sample, 145 RNAs were differentially contained in EVs from mothers of children in the high-risk and low-risk groups, including 31 upregulated and 114 downregulated RNAs in the high-risk group relative to the low-risk group (**Figure 2A, Additional File 1**). In male-carrying pregnancies, we identified 144 differentially EV-contained RNAs (87 upregulated, 57 downregulated), whereas 231 differentially EV-contained RNAs were identified among female-carrying pregnancies (75 upregulated, 156 downregulated) (**Figure 2B, 2C, Additional File 2, Additional File 3**).

**Figure 2.**
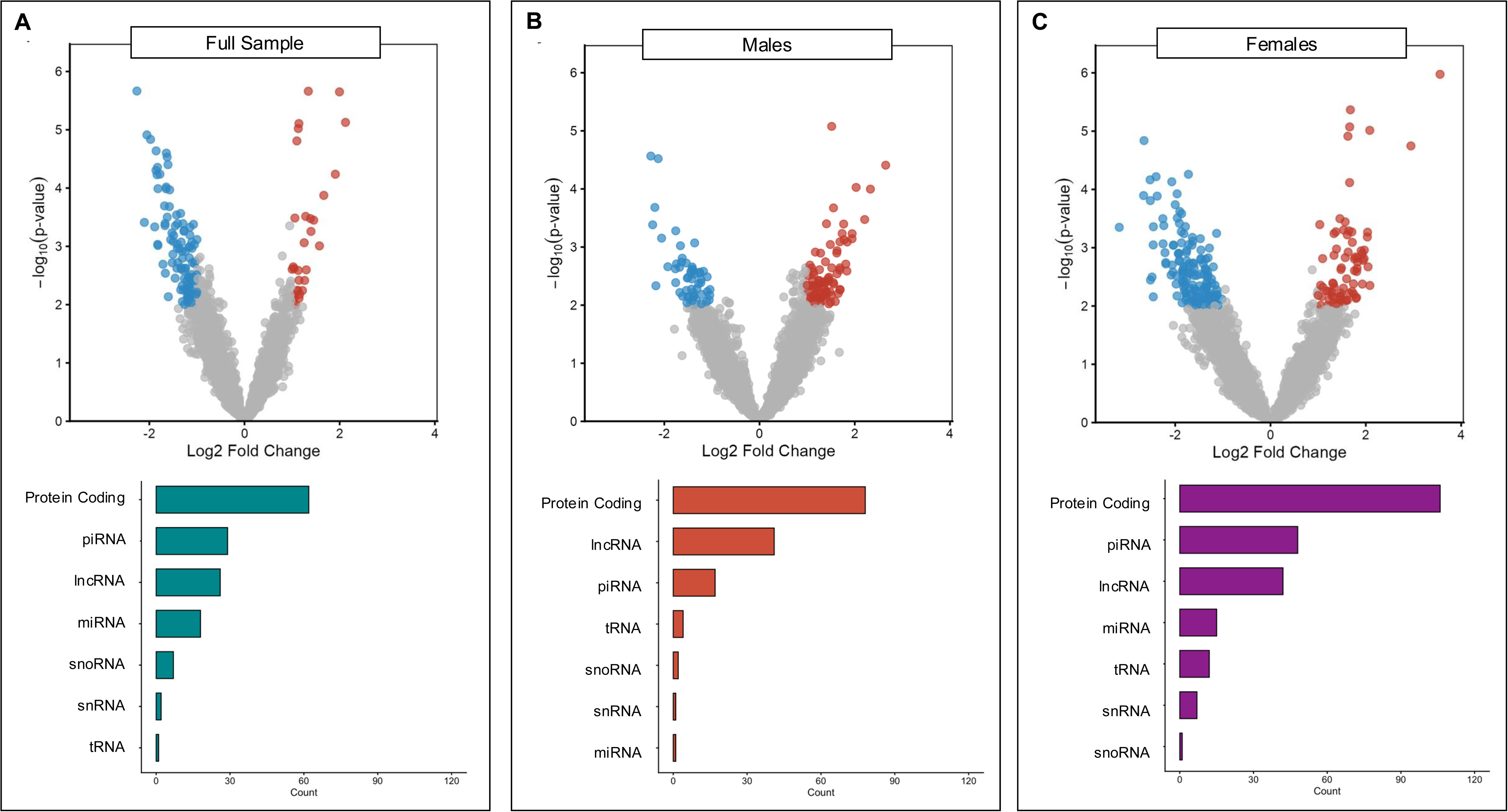
Differentially contained RNA cargo of second trimester maternal plasma EVs associated with autism risk. Volcano plots showing differential representation of RNAs in second-trimester plasma EVs from mothers of children later classified as high autism risk versus low autism risk. The x-axis represents log fold change, and the y-axis represents −log *(p-*value). Points are colored by significance and direction of effect: red indicates significantly upregulated RNAs (log fold change > 1 and *p* < 0.01), blue indicates significantly downregulated RNAs (log fold change < −1 and *p* < 0.01), and gray indicates non-significant transcripts. The distribution of RNA types represented within the significantly differentially contained cargo are shown in the bar graphs below. (A) Full sample including both male and female children. (B) Male children only. (C) Female children only.

In the full sample, the differentially expressed transcripts consisted primarily of protein-coding RNAs (43%), followed by piRNAs (20%), lncRNAs (18%), miRNAs (12%), and smaller proportions of snoRNAs (5%), snRNAs (1%), and tRNAs (1%) (**Figure 2A**) Among male pregnancies, the distribution of RNAs was composed of 54% protein-coding RNAs, 28% lncRNAs, 12% piRNAs, 3% tRNAs, 1% snoRNAs, and 1% miRNAs, and 1% snRNAs, whereas the distribution among the female pregnancies was 46% protein-coding RNAs, 21% piRNAs, 18% lncRNAs,, 6% miRNAs, 5% tRNAs, 3% snRNAs, and 1% snoRNAs (**Figure 2B, 2C**).

The top 10 differentially contained protein-coding RNAs were *PPP3CB, CDH22*, *VEGFC*, *SYNPO*, *TGFBR1*, *APOBR*, *PIGS*, *INKA2*, *N4BP1*, and *ASH2L* (**Table 2**). Enriched KEGG pathways were broadly related to axon guidance, MAPK-mediated signaling, ubiquitin-dependent protein regulation, endocytosis, and cellular stress/inflammatory pathways (**Figure 3A**). The enriched GO Biological Processes terms suggest that the differentially contained EV protein-coding RNAs are associated with developmental and cellular regulatory programs including axonal growth and guidance, proliferative and migratory processes, intracellular signaling, protein turnover, and transcriptional/chromatin organization (**Figure 3B**). Finally, while the SFARI Gene database was not significantly enriched among differentially contained RNAs, 6 of the 62 (9.7%) protein-coding RNAs overlap with genes in the SFARI Gene database, including *AGAP1*, *CDH22*, *CYFIP1*, *PHIP*, *SLC9A9*, and *SSRP1* (**Table 3**).

**Figure 3.**
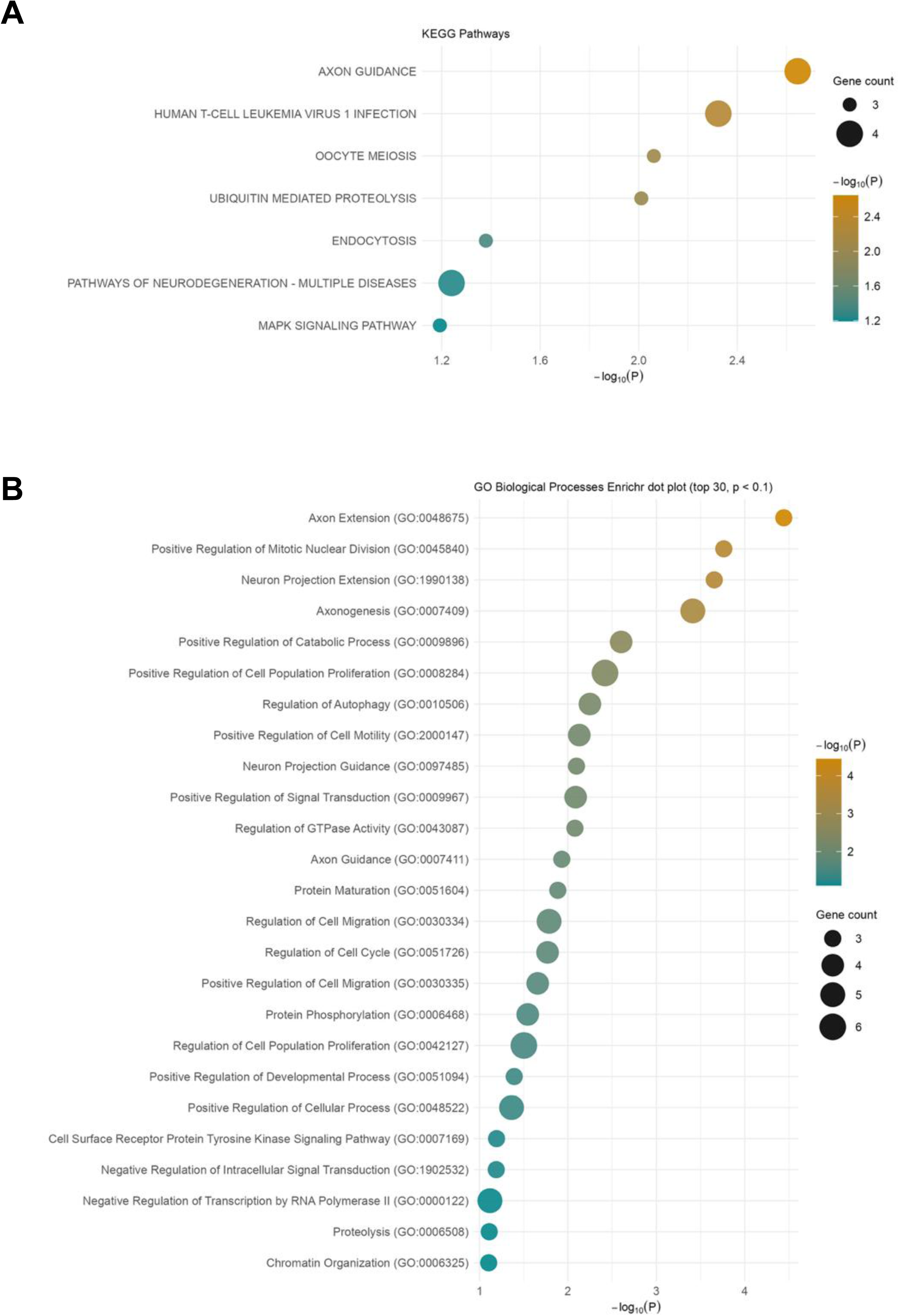
Functional enrichment of protein-coding RNAs differentially represented in second-trimester maternal plasma EVs associated with offspring autism risk. The 62 protein-coding RNAs that were differentially contained in EVs isolated from second-trimester plasma of mothers with children later classified as high risk for autism versus low risk were subjected to pathway and gene ontology enrichment analysis using EnrichR. (A) KEGG pathway enrichment analysis of differentially represented protein-coding RNAs. The dot plot displays the top enriched pathways ranked by nominal P-value. The x-axis represents –log10(P), and dot size reflects the number of genes from the input list contributing to each pathway. Color intensity corresponds to enrichment significance. (B) Gene Ontology (GO) Biological Processes enrichment analysis. The top significantly enriched terms are shown, ranked by nominal P-value. Dot size indicates gene count per term and color represents –log10(P).

**Table 2.**
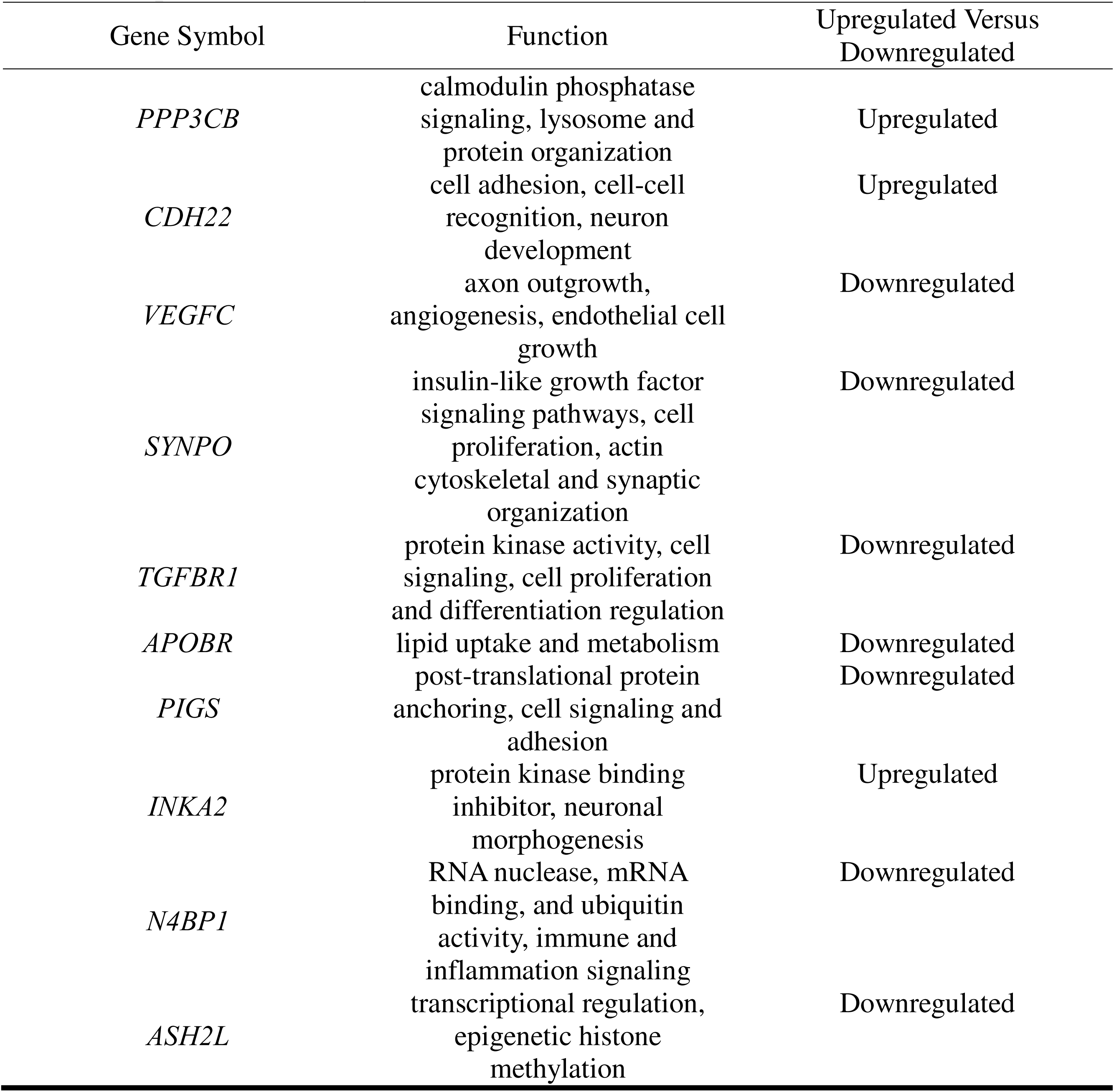
Top 10 Differentially EV-Contained RNAs.

**Table 3.** Differentially EV-Contained RNAs Overlapping with SFARI Gene List.

|  |  |  |  |  |  |
| --- | --- | --- | --- | --- | --- |
| Gene | Chromosome | Genetic Category | SFARI Gene Score | Function | Upregulated Versus Downregulated |
| *AGAP1* | 2 | Rare Single Gene Mutation | 2 | endolysosomal trafficking, reduced axonal terminal size | Downregulated |
| *CDH22* | 20 | Rare Single Gene Mutation, Genetic Association | 2 | cell adhesion | Upregulated |
| *CYFIP1* | 15 | Rare Single Gene Mutation, Genetic Association, Functional | 2 | axon outgrowth | Downregulated |
| *PHIP* | 6 | Rare Single Gene Mutation, Syndromic | 1 | insulin-like growth factor signaling pathways, cell proliferation, cytoskeletal organization | Downregulated |
| *SLC9A9* | 3 | Rare Single Gene Mutation, Genetic Association, Functional | 2 | pH regulation | Downregulated |
| *SSRP1* | 11 | Rare Single Gene Mutation | 2 | nucleosome disassembly, transcription elongation | Downregulated |

## Discussion

Current autism diagnostic practices depend on the observation of behavioral symptoms that usually emerge in toddlerhood or childhood, motivating efforts to identify earlier biological indicators of autism likelihood. Further, understanding biological mechanisms underlying autism may advance targeted therapeutic interventions. Unlike prior studies focused on postnatal samples or mature cell models, we examined EVs during pregnancy from mothers of children who later exhibit greater autism likelihood. Our findings demonstrate some consistency but also divergence from the previous research investigating postnatal EVs and autism.

In contrast to earlier studies reporting both increased and decreased circulating EV concentrations in autistic individuals^20,21^, we found no association between second-trimester maternal plasma EV concentration and total SCQ scores or autism risk. These discrepancies may reflect differences in biospecimen type (serum, plasma), timing of collection (childhood samples, maternal prenatal samples), and EV isolation and characterization methods (ultracentrifugation, size exclusion chromatography (SEC), membrane-based affinity binding kits), all of which can substantially affect measured EV yield and composition^48^. More broadly, the lack of methodological standardization across EV studies remains a major barrier to comparability. Consistent with this, CD63, a canonical EV protein marker, was also not a significant predictor of SCQ scores or autism risk, in line with reports showing null CD63 findings despite differences in other EV features^49,50^.

Small RNA-seq identified differentially EV-contained RNA transcripts enriched for neurodevelopment pathways, including axonogenesis and neuron projection. Since EVs mediate intercellular communication, altered EV cargo may reflect differences in maternal, placental, or fetal signaling environments during critical periods of neurodevelopment during gestation. Notably, many of these transcripts were long non-coding RNAs (lncRNAs), which regulate gene expression and are important for maternal-fetal communication^51^ and nervous system development^52^. The prominence of lncRNAs among our differentially contained RNA transcripts suggests that prenatal EV-associated signaling may involve broad regulatory networks. Additionally, miRNAs may reflect pathways through which EVs could epigenetically mediate exposures and physiological factors within and beyond the intrauterine environment. Most differentially contained RNAs (∼79%) were downregulated, including 7 of the top 10 protein-coding transcripts, consistent with prior EV studies in autism reporting predominantly downregulated RNAs EV cargo^21^.

Protein-coding RNAs comprise a majority of the differentially contained RNAs we identified in the overall sample and in sex-stratified analyses. The top ten differentially contained RNAs were *PPP3CB*, *CDH22*, *VEGFC*, *SYNPO*, *TGFBR1*, *APOBR*, *PIGS*, *INKA2*, *N4BP1*, and *ASH2L*, which converge on several processes strongly implicated in autism and early neurodevelopment, including synaptic plasticity, dendritic spine maturation, axon guidance, cortical development, immune regulation, lipid transport, and embryonic growth. PPP3CB encodes calcineurin, a calmodulin-regulated phosphatase that plays an important role in synaptic transmission and plasticity, and disruptions in calcineurin signaling have been linked to social and behavioral deficits consistent with autism^53–59^. Similarly, CDH22 belongs to the cadherin family of cell adhesion molecules that guide synaptogenesis, axon guidance, dendritic spine regulation, and neural circuit formation, processes repeatedly implicated in autism^60–63^. SYNPO and INKA2 further support this convergence, as both are involved in dendritic spine structure and actin remodeling, suggesting that altered EV-contained RNAs may reflect differences in pathways important for synaptic maturation and neural connectivity^64–68^. Other differentially contained RNAs point to broader developmental, immune, and metabolic mechanisms that may shape neurodevelopmental trajectories. VEGFC is involved in angiogenesis, endothelial cell growth, embryonic development, implantation, and placentation, and also functions as a trophic factor for neural progenitors in the embryonic brain^69–72^. TGFBR1 and ASH2L implicate TGF-β signaling and epigenetic regulation, pathways relevant to microglial function, neurite development, immune regulation, and cortical development^73,74^. TGF-β receptor activity influences axon number and specification during embryonic development, while altered TGF-β signaling has been associated with atypical neurocognitive development and reduced TGF-β1 levels in autistic children^74–82^. ASH2L also supports histone H3K4 methylation and has been linked to embryonic cortical development, postnatal neurogenesis, and intellectual disability^83,84^. In parallel, APOBR highlights the importance of maternal-fetal lipid transport for fetal brain growth^85–88^, while PIGS implicates glycosylphosphatidylinositol biosynthesis, a pathway required for normal embryonic and neural development and associated with autism, developmental delay, seizures, and hypotonia in rare genetic cases^89–94^. Finally, N4BP1 points to inflammatory regulation through NF-κB signaling, which has been reported to be altered in autistic children^95–98^. Collectively, these findings suggest that altered EV RNA cargo during early development may reflect or influence interconnected synaptic, immune, metabolic, and developmental pathways relevant to autism pathogenesis.

### Limitations

Several limitations to our study should be considered. The prospective nuMoM2b study was established to evaluate risk factors for adverse pregnancy outcomes in nulliparous patients, therefore, the study design was not optimized specifically to capture or enrich the study population for women whose children were at high risk of autism. Given the composition of the study population, our analysis aimed to maximize the sample size and included data from all 270 participants who provided maternal plasma during second trimester and who completed parent-report child developmental questionnaires at age 4.5-6 years. The relatively small sample size of the current study may have limited our ability to detect group differences in EV concentration; however, these findings provide compelling preliminary data that can inform and support larger future studies. Additionally, autism likelihood was assessed using a parent-report measure that, although widely used, may capture broader neurodevelopmental differences. Assessment at a single timepoint also limits our ability to account for behavioral features that may emerge later in development. Prenatal sampling was also limited to one timepoint, therefore, assessment whether differences exist across pregnancy is needed. In addition, the origin and target of the maternal plasma EVs remains unknown. The observed transcriptional differences may reflect maternal, fetal, or placental EV populations, and it is not yet known whether these EVs definitively enter the fetal compartment, including the developing brain. If primarily maternal in origin, EV cargo may capture maternal physiological states, including inflammatory states that may emerge prior to the clinical presentation of conditions such as GDM and preeclampsia, that influence fetal neurodevelopment or heritable features that drive similar changes in the fetus. Although fetal EVs are known to enter maternal circulation, it is unclear what proportion of the EVs we captured are fetally-derived EVs. While placentally-derived EVs appear to exert specific effects on target cells and reflect broader placental and gestational health, the shared underlying genetics of the fetus and placenta may drive placental EV packaging, release, and trafficking^33,99^. Therefore, further research is needed to determine the origin, trafficking, and functional impact of these EVs.

An unresolved question is whether EV differences should be considered biomarkers of altered development or active mediators of autism-relevant signaling. Although our findings do not distinguish between these possibilities, they support the potential relevance of EV cargo in neurodevelopmental vulnerability. Our findings should also be considered within a broader bioethical context. The development of prenatal biomarkers raises important scientific, social, and ethical challenges, particularly given the heterogeneity of autism and its complex biology^100^. Current diagnostic practices rely on comprehensive behavioral assessments, which are resource-intensive and not universally accessible; while EV-based biomarkers may offer a complementary approach, they must be developed and interpreted cautiously. Despite current limitations in the field of EV research, EVs continue to be viewed as a promising candidate biomarker for autism, and as a therapeutic platform. Preclinical studies of stem cell-derived EVs suggest they can ameliorate autism-related behaviors and neuroinflammatory changes^101–103^. Although these findings remain preliminary EVs represent a compelling, emerging avenue for autism screening and intervention.

Our study advances EV-based research in autism by providing, to our knowledge, the first evidence of childhood autism risk being associated with altered EV RNA cargo measured during pregnancy. These findings provide a basis for future research aimed at further elucidating the role of prenatal EVs as a biological mechanism, biomarker, and therapeutic delivery system for autism and related neurodevelopmental conditions.

## Data Availability

All data produced in the present study are available upon reasonable request to the authors. Some data are available online at https://dash.nichd.nih.gov/

## Declarations

### Ethics approval and consent to participate

The Columbia University Institutional Review Board reviewed and approved all study procedures (AAAQ7665). Informed written consent was obtained from all participants. Participants included in this analysis provided additional consent for future use of data and future contact by research staff.

### Consent for publication

Not applicable.

### Availability of data and materials

The nuMoM2b Study data are available through the NICHD Data and Specimen Hub (DASH), subject to DASH data access procedures and approval. Data generated for the present analysis are available from the corresponding author upon reasonable request and in accordance with institutional approvals and participant consent.

## Author Contributions

Conceptualization: MF, RW, DH, WG, RS, UR

Formal analysis: MF, WY, SN, DM, CV

Investigation: MF, WY, SN, DM, CV, RW

Resources: RW, DH, WG, RS, UR, HL, WF, AP

Data Curation: MF, WY, SN, RW, WG, RS, UR

Writing: MF, DG, WY, SN

Editing: CV, HL, DH, WG, RS, UR, AP

Visualization: MF, DG, WY, SN

Funding acquisition: MF

All authors read and approved the final manuscript.

## Acknowledgments

The authors wish to thank the nuMoM2b Steering Committee and the entire nuMoM2b study team for providing the resources necessary to support this work. The authors also acknowledge Dr. Frances Champagne, who facilitated the acquisition of the biological samples and data included in the analysis, and Dr. Maya Deyssenroth, who provided technical expertise and access to equipment. Additionally, the authors thank Dr. Courtney McDermott, Dr. Rajesh Patel, and the Rutgers Robert Wood Johnson Medical School Core Imaging Laboratory for providing assistance with transmission electron microscopy. Finally, the authors extend particular gratitude to the families enrolled in the nuMoM2b study.

