## Additional File 1 for "Maternal Extracellular Vesicles During Pregnancy and Autism Risk in Children"

| Additional File 1 Differentially EV-Contained RNAs (Full Sample) | | | |
| --- | --- | --- | --- |
| **Gene** | **log2 Fold Change** | ***p*-value** | **Subtype** |
| LINC01819 | -2.26 | 2.16E-06 | lncRNA |
| hsa-miR-7975 | 2.12 | 7.48E-06 | miRNA |
| hsa-miR-382-3p | -2.10 | 3.88E-04 | miRNA |
| piR-hsa-1294500 | -2.05 | 1.23E-05 | piRNA |
| PPP3CB | 2.00 | 2.24E-06 | Protein Coding |
| piR-hsa-2841762 | -1.97 | 1.46E-05 | piRNA |
| CDH22 | 1.91 | 5.79E-05 | Protein Coding |
| piR-hsa-3724356 | -1.89 | 4.64E-04 | piRNA |
| piR-hsa-1237353 | -1.86 | 4.97E-05 | piRNA |
| SNORD3B-1 | -1.86 | 2.30E-05 | snoRNA |
| hsa-miR-410-3p | -1.84 | 5.90E-05 | miRNA |
| LINC02228 | -1.83 | 4.42E-05 | lncRNA |
| VEGFC | -1.82 | 9.26E-04 | Protein Coding |
| piR-hsa-1236630 | -1.82 | 9.66E-04 | piRNA |
| SYNPO | -1.82 | 1.02E-04 | Protein Coding |
| SNORD3B-2 | -1.78 | 5.78E-05 | snoRNA |
| TGFBR1 | -1.71 | 2.03E-03 | Protein Coding |
| SNORD3D | -1.68 | 2.02E-04 | snoRNA |
| APOBR | -1.67 | 4.40E-04 | Protein Coding |
| PIGS | -1.67 | 4.05E-04 | Protein Coding |
| RNU2-15P | 1.67 | 1.33E-04 | snRNA |
| hsa-miR-539-3p | -1.66 | 2.89E-03 | miRNA |
| SNORD3C | -1.65 | 1.03E-04 | snoRNA |
| LINC01521 | -1.65 | 9.60E-05 | lncRNA |
| piR-hsa-2851584 | -1.64 | 2.52E-05 | piRNA |
| piR-hsa-142694 | -1.63 | 2.94E-05 | piRNA |
| piR-hsa-1891781 | -1.62 | 3.16E-04 | piRNA |
| piR-hsa-142803 | -1.61 | 3.97E-05 | piRNA |
| piR-hsa-1235721 | -1.60 | 7.31E-03 | piRNA |
| SNORD3A | -1.58 | 2.08E-04 | snoRNA |
| INKA2 | 1.58 | 9.82E-04 | Protein Coding |
| NCKAP5-AS2 | -1.57 | 1.07E-04 | lncRNA |
| piR-hsa-175708 | -1.55 | 7.69E-04 | piRNA |
| N4BP1 | -1.53 | 5.87E-04 | Protein Coding |
| hsa-miR-145-3p | -1.52 | 1.91E-03 | miRNA |
| hsa-miR-382-5p | -1.50 | 1.37E-03 | miRNA |
| hsa-miR-485-3p | -1.50 | 8.25E-04 | miRNA |
| ASH2L | -1.49 | 6.49E-04 | Protein Coding |
| LINC01746 | -1.46 | 3.52E-03 | lncRNA |
| ENTREP2 | -1.45 | 4.45E-04 | Protein Coding |
| KCNJ4 | 1.45 | 3.55E-04 | Protein Coding |
| ULK2 | -1.43 | 2.90E-04 | Protein Coding |
| hsa-miR-103a-1-5p | -1.42 | 2.38E-03 | miRNA |
| piR-hsa-2729436 | -1.41 | 9.20E-04 | piRNA |
| CYFIP1 | -1.41 | 1.08E-03 | Protein Coding |
| SEPTIN5 | -1.41 | 4.08E-04 | Protein Coding |
| hsa-miR-376b-3p | -1.40 | 1.90E-03 | miRNA |
| SMIM7 | 1.40 | 5.54E-04 | Protein Coding |
| TERLR1 | 1.39 | 3.34E-04 | lncRNA |
| MIR493HG | -1.36 | 1.10E-03 | lncRNA |
| CHAF1A | -1.34 | 2.72E-04 | Protein Coding |
| piR-hsa-721860 | 1.34 | 2.18E-06 | piRNA |
| piR-hsa-1253722 | -1.34 | 5.57E-03 | piRNA |
| hsa-miR-654-3p | -1.33 | 2.32E-03 | miRNA |
| hsa-miR-487b-3p | -1.33 | 8.43E-04 | miRNA |
| SNAP23 | -1.33 | 2.45E-03 | Protein Coding |
| PLXDC2 | -1.32 | 1.22E-03 | Protein Coding |
| SIDT1 | -1.31 | 4.74E-03 | Protein Coding |
| hsa-miR-369-3p | -1.31 | 8.86E-03 | miRNA |
| LINC01234 | -1.31 | 1.60E-03 | lncRNA |
| piR-hsa-7720620 | 1.30 | 2.53E-03 | piRNA |
| ANAPC5 | -1.29 | 4.08E-04 | Protein Coding |
| hsa-miR-543 | -1.28 | 2.59E-03 | miRNA |
| SH3PXD2A | 1.28 | 3.06E-04 | Protein Coding |
| LINC01667 | -1.28 | 5.62E-03 | lncRNA |
| PDE3A | -1.27 | 5.52E-04 | Protein Coding |
| RNU2-2P | -1.27 | 5.34E-04 | snRNA |
| SALRNA2 | 1.26 | 3.85E-03 | lncRNA |
| MYO1C | -1.26 | 3.59E-03 | Protein Coding |
| SNORD14E | -1.26 | 9.95E-03 | snoRNA |
| piR-hsa-725044 | 1.25 | 8.67E-04 | piRNA |
| SSRP1 | -1.24 | 2.40E-03 | Protein Coding |
| piR-hsa-2857749 | -1.24 | 1.22E-03 | piRNA |
| ZEB1-AS1 | -1.24 | 2.38E-03 | lncRNA |
| SLMAP | -1.23 | 5.39E-03 | Protein Coding |
| GGTA1 | -1.22 | 5.56E-03 | Protein Coding |
| LINC02631 | 1.22 | 5.72E-03 | lncRNA |
| hsa-miR-136-3p | -1.21 | 3.93E-03 | miRNA |
| DDX11L16 | -1.21 | 5.56E-03 | lncRNA |
| SEMA4F | -1.21 | 4.66E-03 | Protein Coding |
| piR-hsa-2828071 | -1.20 | 6.18E-03 | piRNA |
| SNORA61 | -1.20 | 7.29E-03 | snoRNA |
| piR-hsa-2844725 | -1.20 | 8.91E-03 | piRNA |
| LINC02402 | -1.20 | 7.91E-03 | lncRNA |
| LINC00173 | -1.19 | 8.55E-03 | lncRNA |
| hsa-miR-381-3p | -1.19 | 8.56E-03 | miRNA |
| TRDN-AS1 | -1.17 | 1.03E-03 | lncRNA |
| NOP53 | -1.17 | 3.33E-03 | Protein Coding |
| piR-hsa-166257 | -1.17 | 3.41E-03 | piRNA |
| GRK3 | -1.17 | 7.65E-03 | Protein Coding |
| ARHGAP11B-DT | -1.16 | 1.57E-03 | lncRNA |
| KHSRP | -1.16 | 5.06E-03 | Protein Coding |
| BACE2 | 1.16 | 6.67E-03 | Protein Coding |
| CAPNS1 | -1.15 | 6.29E-03 | Protein Coding |
| hsa-miR-127-3p | -1.15 | 6.39E-04 | miRNA |
| piR-hsa-2804856 | -1.15 | 7.64E-03 | piRNA |
| ZSCAN5A-AS1 | -1.15 | 1.01E-03 | lncRNA |
| RABGAP1 | -1.15 | 4.70E-04 | Protein Coding |
| DGCR11 | 1.15 | 7.79E-03 | lncRNA |
| ZC3H18 | 1.15 | 3.82E-03 | Protein Coding |
| piR-hsa-1303866 | 1.14 | 7.81E-06 | piRNA |
| SOX1 | -1.14 | 9.11E-03 | Protein Coding |
| CDC20 | -1.14 | 7.73E-03 | Protein Coding |
| piR-hsa-1250731 | 1.13 | 9.52E-06 | piRNA |
| SORL1 | 1.13 | 2.58E-03 | Protein Coding |
| CCDC82 | -1.13 | 4.99E-03 | Protein Coding |
| ARHGEF2-AS2 | 1.12 | 5.69E-03 | lncRNA |
| piR-hsa-2858643 | -1.12 | 9.16E-03 | piRNA |
| HSBP1 | -1.12 | 4.37E-03 | Protein Coding |
| ATP6V0A1 | -1.11 | 3.04E-03 | Protein Coding |
| LSM14A | -1.11 | 9.53E-03 | Protein Coding |
| ZNF324 | 1.11 | 5.74E-03 | Protein Coding |
| hsa-miR-22-5p | -1.11 | 3.85E-03 | miRNA |
| PRDM6-AS1 | -1.11 | 8.49E-04 | lncRNA |
| SPNS3 | -1.11 | 1.08E-03 | Protein Coding |
| piR-hsa-2854993 | 1.10 | 1.55E-05 | piRNA |
| LRCH1 | -1.10 | 6.51E-03 | Protein Coding |
| ZFR | -1.10 | 1.90E-03 | Protein Coding |
| UBC | 1.08 | 9.97E-03 | Protein Coding |
| Homo_sapiens_tRNA-Ser-GCT-4_5p | -1.07 | 4.19E-04 | tRNA |
| NOPCHAP1 | -1.07 | 3.36E-03 | Protein Coding |
| NAMPT-AS1 | -1.07 | 6.69E-03 | lncRNA |
| piR-hsa-2845517 | 1.06 | 3.27E-04 | piRNA |
| ZNF222-DT | -1.06 | 7.91E-03 | lncRNA |
| PCGF3 | -1.05 | 7.19E-03 | Protein Coding |
| UNC13C | -1.05 | 3.89E-03 | Protein Coding |
| RPA1 | -1.05 | 8.68E-03 | Protein Coding |
| PHIP | -1.05 | 1.98E-03 | Protein Coding |
| piR-hsa-802409 | -1.05 | 9.29E-04 | piRNA |
| SH3BP4 | 1.05 | 7.58E-03 | Protein Coding |
| UNC80 | 1.04 | 2.41E-03 | Protein Coding |
| FERMT3 | -1.04 | 8.10E-03 | Protein Coding |
| INF2 | -1.03 | 1.82E-03 | Protein Coding |
| POLR3C | 1.03 | 2.25E-03 | Protein Coding |
| NEPRO-AS1 | 1.02 | 7.13E-03 | lncRNA |
| EPHB2 | 1.02 | 9.09E-03 | Protein Coding |
| ZFPM2 | -1.02 | 9.80E-03 | Protein Coding |
| CTBP1-DT | -1.02 | 3.94E-03 | lncRNA |
| piR-hsa-758078 | -1.01 | 6.59E-03 | piRNA |
| AGAP1 | -1.01 | 8.92E-03 | Protein Coding |
| SLC9A9 | -1.01 | 3.08E-03 | Protein Coding |
| hsa-miR-146a-5p | -1.00 | 9.12E-03 | miRNA |
| piR-hsa-2867014 | -1.00 | 6.09E-03 | piRNA |
| PTCH1 | 1.00 | 2.47E-03 | Protein Coding |
| NORAD | -1.00 | 7.65E-04 | lncRNA |
