## Additional File 2 for "Maternal Extracellular Vesicles During Pregnancy and Autism Risk in Children"

| Additional File 2: Differentially EV-Contained RNAs (Males) | | | |
| --- | --- | --- | --- |
| **Gene** | **log2 Fold Change** | ***p*-value** | **Subtype** |
| hsa-miR-7975 | 2.65 | 3.90E-05 | miRNA |
| KCNJ4 | 2.33 | 1.01E-04 | Protein Coding |
| LINC01819 | -2.28 | 2.72E-05 | lncRNA |
| SIDT1 | -2.24 | 4.14E-04 | Protein Coding |
| piR-hsa-782835 | 2.21 | 3.36E-04 | piRNA |
| N4BP1 | -2.20 | 2.08E-04 | Protein Coding |
| VEGFC | -2.18 | 4.64E-03 | Protein Coding |
| ENTREP2 | -2.13 | 3.01E-05 | Protein Coding |
| NCKAP5-AS2 | -2.06 | 7.03E-04 | lncRNA |
| POU2F1 | 2.03 | 9.40E-05 | Protein Coding |
| LINC02631 | 1.94 | 5.89E-04 | lncRNA |
| piR-hsa-2851584 | 1.94 | 7.20E-04 | piRNA |
| SYNPO | -1.93 | 2.18E-03 | Protein Coding |
| ARL3 | 1.84 | 8.18E-04 | Protein Coding |
| HECA | 1.83 | 2.59E-03 | Protein Coding |
| PAG1 | 1.81 | 1.96E-03 | Protein Coding |
| piR-hsa-2864966 | 1.78 | 7.51E-04 | piRNA |
| LINC00941 | 1.77 | 4.02E-04 | lncRNA |
| piR-hsa-183121 | -1.77 | 2.37E-03 | piRNA |
| LRP3 | -1.76 | 4.10E-03 | Protein Coding |
| CHAF1A | -1.76 | 5.26E-04 | Protein Coding |
| piR-hsa-779798 | -1.75 | 1.95E-03 | piRNA |
| STAT5A | 1.74 | 5.84E-04 | Protein Coding |
| CLMAT3 | 1.69 | 5.47E-03 | lncRNA |
| CPNE9 | 1.69 | 5.36E-03 | Protein Coding |
| ALK | 1.69 | 2.27E-03 | Protein Coding |
| NRSN2 | 1.69 | 8.12E-04 | Protein Coding |
| DGCR11 | 1.69 | 2.48E-03 | lncRNA |
| NIN | -1.68 | 2.23E-03 | Protein Coding |
| FKTN | -1.67 | 6.06E-03 | Protein Coding |
| ITPR3 | -1.66 | 9.49E-04 | Protein Coding |
| piR-hsa-2865780 | 1.65 | 6.28E-03 | piRNA |
| ANAPC5 | -1.64 | 1.81E-03 | Protein Coding |
| CLDN2 | 1.63 | 1.16E-03 | Protein Coding |
| CUL7 | 1.63 | 2.89E-03 | Protein Coding |
| NUP133-DT | -1.62 | 1.55E-03 | lncRNA |
| LINC01202 | 1.62 | 1.27E-03 | lncRNA |
| piR-hsa-1237353 | -1.62 | 7.01E-03 | piRNA |
| KAT6B | 1.59 | 8.66E-03 | Protein Coding |
| GLIS3 | 1.56 | 4.33E-03 | Protein Coding |
| PDIK1L | 1.55 | 2.12E-04 | Protein Coding |
| Homo_sapiens_tRNA-Gln-CTG-5_5p | 1.55 | 4.09E-03 | tRNA |
| FBN3 | -1.54 | 1.88E-03 | Protein Coding |
| RELL2 | -1.54 | 5.57E-03 | Protein Coding |
| piR-hsa-2777421 | -1.53 | 9.08E-03 | piRNA |
| piR-hsa-2695542 | -1.52 | 3.42E-03 | piRNA |
| SPAG9 | 1.52 | 2.28E-03 | Protein Coding |
| LIG3 | 1.52 | 8.38E-06 | Protein Coding |
| NFE4 | 1.51 | 6.82E-03 | lncRNA |
| piR-hsa-80428 | -1.51 | 7.23E-03 | piRNA |
| ATP6V0A1 | -1.50 | 3.33E-03 | Protein Coding |
| UBE2O | 1.49 | 8.89E-03 | Protein Coding |
| KCNH3 | -1.49 | 7.20E-03 | Protein Coding |
| TARID | 1.49 | 9.04E-04 | lncRNA |
| MAP3K15 | 1.48 | 5.86E-03 | Protein Coding |
| LINC02999 | 1.46 | 3.31E-03 | lncRNA |
| piR-hsa-7720620 | 1.46 | 9.51E-03 | piRNA |
| LINC01087 | 1.45 | 3.79E-03 | lncRNA |
| APPBP2-DT | -1.45 | 9.03E-03 | lncRNA |
| SEC24B-AS1 | -1.44 | 2.63E-03 | lncRNA |
| SSTR4 | 1.44 | 3.47E-03 | Protein Coding |
| SLIT1 | -1.44 | 2.80E-03 | Protein Coding |
| IKZF1 | -1.43 | 5.30E-03 | Protein Coding |
| ATF2 | -1.42 | 3.54E-03 | Protein Coding |
| ZSCAN5A-AS1 | -1.42 | 2.20E-03 | lncRNA |
| TWF2 | -1.41 | 8.33E-03 | Protein Coding |
| TRDN-AS1 | -1.41 | 2.38E-03 | lncRNA |
| LINC01733 | 1.41 | 5.84E-03 | lncRNA |
| PDLIM2 | 1.41 | 3.98E-04 | Protein Coding |
| CHST3 | -1.40 | 8.89E-03 | Protein Coding |
| HCG22 | -1.40 | 2.49E-03 | lncRNA |
| PAICS | 1.40 | 7.24E-03 | Protein Coding |
| SPRY4-AS1 | 1.39 | 4.09E-03 | lncRNA |
| FUBP3 | -1.39 | 6.07E-03 | Protein Coding |
| piR-hsa-790167 | -1.39 | 7.08E-03 | piRNA |
| ABCA12 | 1.39 | 5.87E-03 | Protein Coding |
| LINC01877 | 1.39 | 1.79E-03 | lncRNA |
| SHOX | -1.38 | 6.98E-03 | Protein Coding |
| HP1BP3 | -1.38 | 9.99E-03 | Protein Coding |
| RABGAP1 | -1.37 | 2.92E-03 | Protein Coding |
| CHAF1B | 1.36 | 6.84E-03 | Protein Coding |
| NSD1 | -1.36 | 8.49E-04 | Protein Coding |
| ADNP-AS1 | 1.36 | 4.16E-03 | lncRNA |
| ZMIZ1-AS1 | 1.36 | 5.12E-03 | lncRNA |
| PPP3CB | 1.35 | 3.96E-03 | Protein Coding |
| LINC01258 | 1.34 | 6.08E-03 | lncRNA |
| FBXW2 | -1.33 | 9.94E-03 | Protein Coding |
| SLC10A1 | 1.33 | 4.14E-03 | Protein Coding |
| GUSBP1 | 1.33 | 6.96E-03 | lncRNA |
| GAPVD1 | -1.33 | 8.28E-03 | Protein Coding |
| SNORA65 | -1.31 | 5.20E-03 | snoRNA |
| FFAR4 | 1.29 | 7.47E-03 | Protein Coding |
| CDV3 | 1.28 | 1.27E-03 | Protein Coding |
| FAM169A | 1.28 | 6.49E-03 | Protein Coding |
| PGPEP1 | -1.28 | 4.28E-03 | Protein Coding |
| SZT2-AS1 | 1.28 | 7.23E-03 | lncRNA |
| FRG1JP | 1.28 | 9.25E-03 | lncRNA |
| FAM86JP | 1.28 | 8.93E-03 | lncRNA |
| MIR210HG | 1.26 | 2.45E-03 | lncRNA |
| Homo_sapiens_tRNA-Met-CAT-1_5p | -1.26 | 2.70E-03 | tRNA |
| DACT3-AS1 | -1.26 | 2.81E-03 | lncRNA |
| NHSL1-AS1 | 1.26 | 3.77E-03 | lncRNA |
| LOXL1-AS1 | 1.24 | 3.78E-03 | lncRNA |
| DYNLL2 | -1.23 | 4.11E-03 | Protein Coding |
| CLOCK | 1.23 | 4.89E-03 | Protein Coding |
| MLN | 1.23 | 4.21E-03 | Protein Coding |
| ALMS1-IT1 | 1.23 | 4.67E-03 | lncRNA |
| CANX | -1.23 | 3.64E-03 | Protein Coding |
| piR-hsa-785547 | -1.22 | 9.45E-03 | piRNA |
| OVCH1-AS1 | -1.21 | 6.55E-03 | lncRNA |
| ANKH | 1.21 | 7.97E-03 | Protein Coding |
| FRMD5 | 1.19 | 7.61E-03 | Protein Coding |
| RNU105C | 1.19 | 9.73E-03 | snoRNA |
| Homo_sapiens_tRNA-Thr-TGT-2_3p | -1.18 | 2.61E-03 | tRNA |
| MAPK14 | 1.18 | 6.35E-03 | Protein Coding |
| CDK6-AS1 | 1.17 | 5.08E-03 | lncRNA |
| SYT11 | 1.17 | 1.22E-03 | Protein Coding |
| SLC47A2 | 1.16 | 9.00E-03 | Protein Coding |
| piR-hsa-5267278 | 1.16 | 9.14E-03 | piRNA |
| SMARCA5-AS1 | 1.16 | 9.44E-03 | lncRNA |
| MYO1B-AS1 | 1.15 | 6.62E-03 | lncRNA |
| RNU5E-9P | 1.14 | 2.99E-03 | snRNA |
| BTNL9 | 1.14 | 6.88E-03 | Protein Coding |
| CT75 | 1.13 | 2.67E-03 | lncRNA |
| KIF23-AS1 | 1.12 | 4.60E-03 | lncRNA |
| RAB18 | 1.12 | 3.67E-03 | Protein Coding |
| BEAN1 | 1.12 | 8.52E-03 | Protein Coding |
| piR-hsa-2774546 | -1.11 | 8.70E-03 | piRNA |
| COL6A5 | -1.10 | 3.26E-03 | Protein Coding |
| piR-hsa-298988 | -1.10 | 5.83E-03 | piRNA |
| SCN5A | 1.09 | 2.29E-03 | Protein Coding |
| TTC6 | 1.09 | 7.13E-03 | Protein Coding |
| HADHB | 1.07 | 4.63E-03 | Protein Coding |
| LINC00559 | 1.07 | 8.00E-03 | lncRNA |
| EXPH5 | 1.07 | 8.76E-03 | Protein Coding |
| LINC02565 | 1.06 | 4.62E-03 | lncRNA |
| Homo_sapiens_tRNA-Ile-AAT-9_5p | 1.06 | 2.05E-03 | tRNA |
| KRT20 | 1.06 | 9.42E-03 | Protein Coding |
| piR-hsa-167034 | -1.06 | 8.14E-03 | piRNA |
| TMEM40 | -1.05 | 6.71E-03 | Protein Coding |
| LINC01770 | -1.04 | 5.38E-03 | lncRNA |
| POLK | 1.03 | 5.82E-03 | Protein Coding |
| KLRK1 | 1.03 | 3.12E-03 | Protein Coding |
| UNC79 | 1.00 | 4.52E-03 | Protein Coding |
