## Additional File 3 for "Maternal Extracellular Vesicles During Pregnancy and Autism Risk in Children"

| Additional File 3: Differentially EV-Contained RNAs (Females) | | | |
| --- | --- | --- | --- |
| **Gene** | **log2 Fold Change** | ***p*-value** | **Subtype** |
| piR-hsa-788189 | 3.56 | 1.06E-06 | piRNA |
| SPAG6 | -3.17 | 4.47E-04 | Protein Coding |
| piR-hsa-758850 | 2.95 | 1.79E-05 | piRNA |
| NAGS | -2.66 | 1.27E-04 | Protein Coding |
| piR-hsa-45772 | -2.65 | 1.45E-05 | piRNA |
| piR-hsa-2857749 | -2.53 | 6.84E-05 | piRNA |
| piR-hsa-1294500 | -2.52 | 1.56E-04 | piRNA |
| piR-hsa-100383 | -2.52 | 3.58E-03 | piRNA |
| piR-hsa-3724356 | -2.49 | 3.18E-03 | piRNA |
| hsa-miR-369-3p | -2.46 | 8.96E-04 | miRNA |
| hsa-miR-539-3p | -2.46 | 6.96E-03 | miRNA |
| hsa-miR-145-3p | -2.46 | 4.37E-04 | miRNA |
| TMC4 | -2.40 | 6.05E-05 | Protein Coding |
| DENND11 | -2.38 | 1.30E-04 | Protein Coding |
| piR-hsa-142694 | -2.26 | 3.17E-04 | piRNA |
| JHY | -2.25 | 8.57E-04 | Protein Coding |
| piR-hsa-142803 | -2.24 | 4.17E-04 | tRNA |
| MYO1C | -2.21 | 1.22E-03 | Protein Coding |
| hsa-miR-410-3p | -2.20 | 1.80E-03 | miRNA |
| LINC02268 | -2.20 | 1.90E-03 | lncRNA |
| hsa-miR-376b-3p | -2.09 | 2.95E-03 | miRNA |
| SLC22A18AS | 2.08 | 4.45E-03 | lncRNA |
| piR-hsa-721860 | 2.08 | 9.69E-06 | piRNA |
| piR-hsa-92465 | -2.07 | 7.39E-05 | piRNA |
| MIR493HG | -2.06 | 1.98E-03 | lncRNA |
| hsa-miR-103a-1-5p | -2.05 | 2.63E-03 | miRNA |
| COL1A1 | 2.05 | 2.13E-03 | Protein Coding |
| RASGRF2 | -2.04 | 9.13E-04 | Protein Coding |
| PKIB | 2.04 | 5.43E-04 | Protein Coding |
| BEX3 | -2.03 | 8.97E-04 | Protein Coding |
| RASIP1 | 2.03 | 6.55E-04 | Protein Coding |
| CRKL | -2.00 | 1.83E-04 | Protein Coding |
| SLC7A5 | -1.98 | 4.20E-03 | Protein Coding |
| piR-hsa-2841762 | -1.98 | 3.02E-03 | piRNA |
| RNU1-132P | 1.97 | 1.10E-03 | snRNA |
| C10orf67 | 1.96 | 1.54E-03 | Protein Coding |
| hsa-miR-382-3p | -1.96 | 6.72E-03 | miRNA |
| ADGRG1 | -1.96 | 1.19E-04 | Protein Coding |
| MAP3K20 | -1.96 | 3.09E-04 | Protein Coding |
| TTC9 | 1.94 | 1.65E-03 | Protein Coding |
| KHSRP | -1.93 | 1.26E-03 | Protein Coding |
| LINC02555 | -1.93 | 1.27E-03 | lncRNA |
| RPS6KB2-AS1 | 1.93 | 1.26E-03 | lncRNA |
| TLE1 | 1.93 | 4.21E-03 | Protein Coding |
| piR-hsa-120994 | -1.92 | 1.24E-03 | piRNA |
| UFD1 | -1.92 | 1.71E-03 | Protein Coding |
| SNHG28 | -1.91 | 3.81E-04 | lncRNA |
| KCNJ15 | -1.91 | 1.64E-03 | Protein Coding |
| ATR | -1.90 | 2.40E-04 | Protein Coding |
| RNU2-15P | 1.90 | 1.13E-03 | snRNA |
| Homo_sapiens_tRNA-Lys-CTT-11_internal | -1.87 | 5.31E-04 | tRNA |
| LEMD3 | -1.87 | 8.26E-04 | Protein Coding |
| CSNK1G2 | -1.87 | 2.61E-04 | Protein Coding |
| SEMA4F | -1.85 | 8.80E-03 | Protein Coding |
| CASC18 | -1.85 | 2.54E-03 | lncRNA |
| SNORD14E | -1.85 | 3.77E-03 | snoRNA |
| LINC01894 | -1.85 | 3.86E-03 | lncRNA |
| EPSTI1 | -1.84 | 2.97E-03 | Protein Coding |
| hsa-miR-487b-3p | -1.83 | 2.36E-03 | miRNA |
| KCTD3 | 1.83 | 1.48E-03 | Protein Coding |
| LSM14A | -1.83 | 2.70E-03 | Protein Coding |
| RNU1-16P | 1.83 | 1.77E-03 | snRNA |
| hsa-miR-543 | -1.83 | 2.63E-03 | miRNA |
| SMCR8 | -1.82 | 4.63E-03 | Protein Coding |
| DNAH7 | -1.82 | 2.18E-03 | Protein Coding |
| PTGER3 | -1.82 | 5.76E-04 | Protein Coding |
| CDH3-AS1 | 1.82 | 2.26E-03 | lncRNA |
| LMNB1 | 1.81 | 2.56E-03 | Protein Coding |
| LINC02125 | -1.81 | 5.46E-03 | lncRNA |
| GAREM1 | 1.81 | 7.14E-03 | Protein Coding |
| ASH2L | -1.80 | 3.81E-03 | Protein Coding |
| piR-hsa-771673 | 1.79 | 7.40E-03 | piRNA |
| hsa-miR-376c-3p | -1.79 | 4.42E-03 | miRNA |
| LINC01234 | -1.79 | 4.35E-04 | lncRNA |
| MYLK4 | -1.78 | 7.84E-03 | Protein Coding |
| LINC01970 | 1.78 | 1.45E-03 | lncRNA |
| piR-hsa-2844725 | -1.78 | 6.00E-03 | piRNA |
| piR-hsa-1891781 | -1.77 | 6.00E-03 | piRNA |
| PDGFRB | -1.77 | 6.97E-03 | Protein Coding |
| MIR9-2HG | -1.76 | 1.21E-03 | lncRNA |
| BETALINC1 | 1.76 | 2.48E-03 | lncRNA |
| piR-hsa-449521 | 1.76 | 1.56E-03 | piRNA |
| RANBP17 | -1.76 | 3.52E-03 | Protein Coding |
| MDC1-AS1 | -1.76 | 1.11E-03 | lncRNA |
| PCGF3 | -1.76 | 1.67E-03 | Protein Coding |
| piR-hsa-175708 | -1.75 | 2.04E-03 | piRNA |
| piR-hsa-372959 | 1.75 | 6.27E-03 | piRNA |
| RFLNB | 1.74 | 4.17E-03 | Protein Coding |
| LINC01619 | -1.72 | 5.50E-05 | lncRNA |
| Homo_sapiens_tRNA-Lys-CTT-7_internal | -1.72 | 8.84E-04 | tRNA |
| piR-hsa-127794 | 1.72 | 8.09E-04 | piRNA |
| hsa-miR-629-5p | -1.71 | 8.84E-03 | miRNA |
| AK2 | 1.71 | 5.64E-03 | Protein Coding |
| PIK3CD | -1.71 | 2.69E-03 | Protein Coding |
| piR-hsa-2845517 | 1.71 | 5.36E-04 | piRNA |
| FAM27C | -1.70 | 1.88E-03 | lncRNA |
| WTIP | -1.69 | 2.78E-03 | Protein Coding |
| hsa-miR-376a-3p | -1.69 | 4.97E-03 | miRNA |
| piR-hsa-1303866 | 1.68 | 4.31E-06 | piRNA |
| PSMC2 | -1.67 | 6.37E-03 | Protein Coding |
| piR-hsa-1250731 | 1.66 | 8.46E-06 | piRNA |
| piR-hsa-2858950 | 1.66 | 7.64E-05 | piRNA |
| MDN1-AS1 | 1.65 | 6.01E-03 | lncRNA |
| ZNF805 | -1.65 | 2.64E-03 | Protein Coding |
| HEG1 | -1.65 | 8.45E-03 | Protein Coding |
| hsa-miR-500a-3p | -1.65 | 9.04E-04 | miRNA |
| UNC80 | 1.64 | 5.13E-04 | Protein Coding |
| PHIP | -1.64 | 6.66E-04 | Protein Coding |
| INSIG1 | -1.63 | 4.37E-03 | Protein Coding |
| piR-hsa-2838937 | -1.63 | 7.74E-04 | piRNA |
| SH3PXD2A | 1.63 | 1.19E-03 | Protein Coding |
| piR-hsa-2854993 | 1.62 | 1.22E-05 | piRNA |
| TUG1 | -1.62 | 1.83E-03 | Protein Coding |
| ERC2 | 1.61 | 7.98E-03 | Protein Coding |
| piR-hsa-95906 | 1.61 | 6.02E-04 | piRNA |
| CENPE | -1.61 | 1.87E-03 | Protein Coding |
| AKAP12 | -1.61 | 2.99E-03 | Protein Coding |
| PTGIR | -1.61 | 8.14E-03 | Protein Coding |
| hsa-miR-22-5p | -1.60 | 6.95E-03 | miRNA |
| UBE2D3 | -1.60 | 5.07E-03 | Protein Coding |
| CFAP44 | -1.60 | 2.21E-03 | Protein Coding |
| LINC01359 | 1.60 | 3.42E-03 | lncRNA |
| piR-hsa-369181 | 1.59 | 7.98E-03 | piRNA |
| WDFY2 | -1.58 | 5.56E-03 | Protein Coding |
| SSRP1 | -1.57 | 7.67E-03 | Protein Coding |
| GAB2 | 1.57 | 1.48E-03 | Protein Coding |
| CCDC88C | 1.57 | 3.60E-04 | Protein Coding |
| LINC01920 | -1.57 | 8.75E-03 | lncRNA |
| LINC02306 | -1.57 | 9.71E-03 | lncRNA |
| MYOZ3-AS1 | -1.56 | 8.10E-03 | lncRNA |
| BAP1 | 1.55 | 8.30E-03 | Protein Coding |
| OR6C4 | -1.55 | 3.01E-03 | Protein Coding |
| NIFK-AS1 | -1.55 | 8.41E-03 | lncRNA |
| NRG1 | -1.54 | 5.50E-03 | Protein Coding |
| CHASERR | -1.54 | 7.23E-03 | lncRNA |
| LINC02288 | 1.53 | 4.96E-04 | lncRNA |
| piR-hsa-753878 | 1.53 | 4.94E-03 | piRNA |
| BTG1 | -1.51 | 9.01E-03 | Protein Coding |
| KCNIP2-AS1 | -1.51 | 9.32E-03 | lncRNA |
| TDRD9 | -1.51 | 8.58E-03 | Protein Coding |
| CD34 | -1.51 | 6.95E-03 | Protein Coding |
| GTPBP2 | -1.49 | 9.22E-03 | Protein Coding |
| piR-hsa-2828222 | 1.49 | 2.22E-03 | piRNA |
| piR-hsa-784964 | -1.49 | 4.51E-03 | piRNA |
| BLTP1 | -1.48 | 2.86E-03 | Protein Coding |
| piR-hsa-193515 | 1.48 | 2.37E-03 | piRNA |
| RNU6-13P | -1.48 | 8.37E-03 | snRNA |
| AP3D1 | 1.47 | 5.98E-03 | Protein Coding |
| GSN | 1.47 | 6.90E-03 | Protein Coding |
| Homo_sapiens_tRNA-Gly-GCC-3_3p | -1.47 | 7.00E-04 | tRNA |
| piR-hsa-52699 | 1.47 | 6.74E-03 | piRNA |
| FKBP8 | 1.47 | 7.87E-03 | Protein Coding |
| SPNS3 | -1.46 | 1.60E-03 | Protein Coding |
| PRDM6-AS1 | -1.46 | 1.54E-03 | lncRNA |
| FAF1 | -1.46 | 1.10E-03 | Protein Coding |
| LINC01134 | 1.46 | 3.18E-04 | lncRNA |
| NALCN-AS1 | 1.44 | 9.09E-03 | lncRNA |
| CD99 | -1.43 | 7.76E-03 | Protein Coding |
| CAMKV | -1.41 | 1.87E-03 | Protein Coding |
| LINC00293 | -1.41 | 6.64E-03 | lncRNA |
| CDC42 | -1.40 | 5.27E-03 | Protein Coding |
| BIRC8 | 1.39 | 9.92E-03 | lncRNA |
| NR3C1 | -1.39 | 2.89E-03 | Protein Coding |
| piR-hsa-177501 | -1.39 | 9.52E-03 | piRNA |
| hsa-miR-127-3p | -1.39 | 4.78E-03 | miRNA |
| ZNF217 | 1.38 | 9.11E-03 | Protein Coding |
| Homo_sapiens_tRNA-Lys-TTT-3_3p | 1.38 | 1.05E-03 | tRNA |
| PSME4 | -1.38 | 7.06E-03 | Protein Coding |
| LINC02202 | 1.37 | 5.33E-04 | lncRNA |
| SIAE | -1.37 | 2.26E-03 | Protein Coding |
| WRNIP1 | -1.36 | 2.38E-03 | Protein Coding |
| NOP53 | -1.36 | 7.77E-03 | Protein Coding |
| FENDRR | -1.36 | 1.58E-03 | lncRNA |
| SRGAP3 | -1.36 | 5.06E-03 | Protein Coding |
| CENPB | -1.35 | 9.45E-03 | Protein Coding |
| SMC6 | -1.35 | 4.81E-03 | Protein Coding |
| ABCB6 | -1.34 | 3.97E-03 | Protein Coding |
| piR-hsa-96321 | -1.34 | 4.79E-03 | piRNA |
| piR-hsa-779798 | 1.34 | 4.75E-03 | piRNA |
| KDM3B | 1.33 | 5.73E-04 | Protein Coding |
| piR-hsa-2840023 | 1.33 | 1.35E-03 | piRNA |
| piR-hsa-99885 | 1.33 | 7.00E-03 | piRNA |
| SNRNP200 | -1.33 | 4.28E-03 | Protein Coding |
| ADAR | -1.33 | 3.19E-03 | Protein Coding |
| EEIG1 | -1.32 | 7.81E-03 | Protein Coding |
| IQCE | -1.32 | 2.55E-03 | Protein Coding |
| RNU1-11P | 1.32 | 9.07E-03 | snRNA |
| piR-hsa-165419 | 1.32 | 4.16E-03 | piRNA |
| CDV3 | -1.31 | 2.96E-03 | Protein Coding |
| EXOC2 | -1.31 | 1.49E-03 | Protein Coding |
| piR-hsa-81900 | 1.31 | 4.03E-03 | piRNA |
| Homo_sapiens_tRNA-Cys-GCA-17_3p | 1.30 | 6.82E-03 | tRNA |
| LINC01394 | -1.30 | 6.61E-03 | lncRNA |
| KLHL7-DT | -1.29 | 6.91E-03 | lncRNA |
| piR-hsa-784665 | 1.29 | 5.80E-03 | piRNA |
| MTNR1A | -1.28 | 1.01E-03 | Protein Coding |
| piR-hsa-441878 | -1.27 | 9.37E-03 | piRNA |
| RIPOR3 | -1.27 | 8.60E-03 | Protein Coding |
| ROCK1P1 | -1.27 | 8.49E-03 | lncRNA |
| AFF1 | -1.25 | 7.57E-03 | Protein Coding |
| PFKL | -1.25 | 6.56E-03 | Protein Coding |
| LINC01141 | -1.24 | 6.10E-03 | lncRNA |
| KLRK1 | -1.24 | 3.43E-03 | Protein Coding |
| piR-hsa-2669041 | 1.23 | 5.70E-03 | piRNA |
| piR-hsa-2705644 | 1.23 | 7.86E-03 | piRNA |
| RNU2-2P | -1.23 | 4.03E-03 | snRNA |
| SPTB | -1.20 | 9.03E-03 | Protein Coding |
| CDK2AP2 | 1.20 | 9.71E-03 | Protein Coding |
| piR-hsa-55506 | -1.20 | 6.58E-03 | piRNA |
| MAGI3 | -1.20 | 2.13E-03 | Protein Coding |
| Homo_sapiens_tRNA-Leu-TAG-1_3p | 1.19 | 5.50E-03 | tRNA |
| KDR | -1.18 | 7.11E-03 | Protein Coding |
| ATP9A | -1.17 | 9.64E-03 | Protein Coding |
| Homo_sapiens_tRNA-Ser-GCT-2_3p | -1.17 | 4.38E-03 | tRNA |
| RNU6-12P | -1.17 | 8.99E-03 | snRNA |
| Homo_sapiens_tRNA-Val-CAC-4_3p | -1.16 | 9.32E-03 | tRNA |
| PICART1 | -1.15 | 6.90E-03 | lncRNA |
| Homo_sapiens_tRNA-Leu-AAG-4_5p | -1.15 | 7.04E-03 | tRNA |
| piR-hsa-585508 | -1.14 | 6.47E-03 | piRNA |
| XPA | -1.13 | 5.67E-04 | Protein Coding |
| CFLAR | -1.13 | 5.30E-03 | Protein Coding |
| LINC00261 | -1.11 | 6.32E-03 | lncRNA |
| piR-hsa-128214 | -1.10 | 9.69E-03 | piRNA |
| VLDLR | 1.10 | 4.78E-03 | Protein Coding |
| Homo_sapiens_tRNA-Glu-TTC-4_5p | 1.09 | 1.54E-03 | tRNA |
| FMNL1-DT | 1.08 | 6.87E-03 | lncRNA |
| LINC01132 | 1.05 | 9.70E-03 | lncRNA |
| Homo_sapiens_tRNA-Glu-TTC-3_5p | 1.05 | 4.42E-03 | tRNA |
| Homo_sapiens_tRNA-Ala-CGC-2_internal | 1.03 | 4.01E-04 | tRNA |
| LINC02575 | -1.02 | 7.61E-03 | lncRNA |
| piR-hsa-1276579 | 1.00 | 6.61E-03 | piRNA |
